# NEUTROPHIL ELASTASE AS A PREDICTIVE BIOMARKER FOR POST-STROKE INFECTION

**DOI:** 10.1101/2025.02.04.25321701

**Authors:** B Díaz-Benito, P Calleja, L Alzamora, A Ruiz, A Martínez-Salio, F Ostos, M Gutiérrez-Sánchez, A García-Culebras, MA Moro, A Moraga, I Lizasoain

**Affiliations:** Neurovascular Research Unit. Pharmacology Department, Complutense Medical School, Instituto Investigación Hospital 12 Octubre (imas12), Madrid, Spain; Department of Neurology and Stroke Center, Hospital 12 Octubre (imas12), Madrid, Spain; Radiology Department, Massachusetts General Hospital, Harvard Medical School, Boston, USA; Neurovascular Research Unit. Cellular Biology and Histology Department, Complutense Medical School, Instituto Investigación Hospital 12 Octubre (imas12), Madrid, Spain; Neurovascular Pathophysiology, Centro Nacional Investigaciones Cardiovasculares (CNIC), Madrid, Spain

## Abstract

**BACKGROUND.:** Ischemic stroke, a major cause of death and disability, triggers immune suppression and inflammation, increasing the risk of infections, particularly early post-stroke. These infections worsen outcomes, prolong hospitalization, and lack reliable predictive protocols. Current management with broad-spectrum antibiotics is not proven effective and may disrupt microbiota balance. Neutrophil elastase (NE), vital for immune defense, can exacerbate inflammation and infection susceptibility. The main objective of this study is to explore the role of NE in the development of infections after ischemic stroke.

**METHODS.:** This study enrolled 544 ischemic stroke patients to evaluate NE as a biomarker for post-stroke infections. Data regarding age, sex, cardiovascular risk factors, stroke etiology and severity were collected. Blood samples from eligible patients, meeting strict inclusion and exclusion criteria, were analyzed using ELISA and multiplex assays to measure NE, myeloperoxidase, and immune mediators.

**RESULTS.:** This study analyzed post-stroke infections in ischemic stroke patients, with 27.57% developing infections, respiratory (48.9%) and urinary (24.8%), mostly within three days. Infected patients were older, more often women, and diabetic, with higher NIHSS scores and larger infarct volumes. Elevated NE levels, higher leukocyte counts, and reduced lymphocytes at admission correlated with infection. Logistic regression showed NE, NIHSS, and diabetes as independent predictors. A matched case-control analysis confirmed higher NE levels in infected cases. Cytokine analysis revealed distinct immune profiles, with NE and IL-6 linked to respiratory infections.

**CONCLUSIONS.:** NE levels at admission may help identify stroke patients at high infection risk, guiding targeted treatment and supporting potential anti-NE therapies and prophylactic antibiotic strategies.

## INTRODUCTION

Ischemic stroke is a devastating disease caused by the obstruction of cerebral blood flow due to thrombus formation. The limited therapeutic options for this disease make it the second leading cause of death worldwide and the primary cause of long-term disability.^1^

Ischemic stroke is characterized by exacerbated inflammation that compromises the immune system, increasing the risk of bacterial infections. These infections, particularly in the first days after a stroke, are associated with poorer outcomes and represent one of the leading life-threatening complications during the subacute phase.^2,3^ Additionally, post-stroke in-hospital infections prolong hospitalization and significantly impact recovery.^2,4^ Identifying patients at risk of infections due to stroke-induced immunosuppression is crucial for improving outcomes.

Currently, most strategies for managing post-stroke infections are based on broad-spectrum antibiotics, either to treat established infections or as prophylactic therapy. However, the effectiveness of this practice is not proven,^5^ as it can disrupt the natural balance of human bacterial communities, causing intestinal microbiota dysbiosis, which may influence infection rates and stroke outcomes.^6,7^ Infection risk is influenced by multiple factors, including age, sex, comorbidities and post-stroke clinical variables such as the use of invasive devices during surgical procedures,^8^ lymphopenia, NIHSS score, and infarct volume.^9,10^ Despite the identification of these risk factors, no established protocols exist to reliably predict or prevent post-stroke infections. Consequently, current guidelines for acute stroke management do not recommend the routine use of antibiotics for infection prevention.^11^

Neutrophils, the most abundant circulating leukocytes, serve as the first line of innate immune defense.^12^ They combat microbial attacks through various molecular and cellular strategies, including phagocytosis, the intracellular generation of reactive oxygen species, degranulation,^13^ and the formation of neutrophil extracellular traps (NETs),^14,15^ which are large fibrous networks of extracellular DNA formed by decondensed chromatin associated with histones and neutrophil granule proteins such as myeloperoxidase (MPO) and neutrophil elastase (NE).^14^

NE is a key component of the immune response and, in addition to participating in microbial elimination through the formation of NETs, it also carries out other mechanisms such as leukocyte transmigration and increasing the release of cytokines. However, dysregulated NE activity can contribute to prolonged inflammation, tissue damage, and weakening of the innate immune system.^16^ Understanding the role of NE is crucial for elucidating its roles in various disease states, including susceptibility to bacterial infections in inflammation-associated conditions such as stroke.

With this background, identifying patients at risk of infection following a stroke is crucial for improving disease outcomes. In our research, we explored the role of NE in the development of infections after ischemic stroke and found an association between higher NE levels upon admission and the development of poststroke infections.

## MATERIAL AND METHODS

### Study design

A total of 544 patients were recruited from the stroke unit at 12 de Octubre Hospital between January 2018 and January 2024, meeting the specified inclusion and exclusion criteria. Of these, 61 patients had missing 90-day modified Rankin Scale (mRS) data. The missing values were considered random and non-systematic, attributed to factors such as missed appointments, transfers to other centers, or incomplete documentation. All samples were derived from patients providing informed consent to participate in this study, which had been formally approved by the ethics committee of the 12 de Octubre Hospital.

Due to the lack of prior research on NE as a biomarker for post-stroke infections, formal sample size calculations were not feasible. However, a preliminary study suggested that around 500 patients would be adequate to identify significant associations. The final sample size was determined pragmatically, based on the number of eligible patients available during recruitment.

### Inclusion criteria

Adults suffering ischemic stroke over 18 years-old; previously independent (pre-morbid mRS ≤2); admitted within 9 hours of symptom-onset/symptom-discovery in wake-up stroke.

### Exclusion criteria

Transient ischemic attacks; lacunar strokes; intracranial bleed secondary to trauma or subarachnoid hemorrhage; previous stroke/acute myocardial infarction/severe systemic infection/major surgery within the last 3 months; active systemic inflammatory disease; immunosuppressive treatment, pregnancy, or puerperium, and underlying active neoplastic disease.

Data was retrieved regarding age, sex, cardiovascular risk factors (CVRF), previous medication and baseline stroke severity (National Institutes of Health Stroke Scale (NIHSS)) (Table 1). EDTA-treated-whole-venous-blood samples were obtained upon admission and in the following 12-24 hours when at the Stroke Unit. Laboratory data from blood tests (biochemistry, cell counts and coagulation) was obtained from the same samples (Table 2). Plasma was extracted and used to quantify different immune response mediators.

**Table 1.** Baseline of demographic and clinical characteristics of ischemic stroke cohort (n=544). Results are expressed in mean (SD) or number of cases (%). P-value in bold are significant (p<0.05).

| n | Post-Stroke Infection in Ischemic Stroke Cohort |  |  |
| --- | --- | --- | --- |
|  | No<br>394 | Yes<br>150 | p |
| Age, mean (SD) | 72.36 (14.41) | 75.25 (13.78) | **0.035** |
| Gender (Female), n (%) | 193 (49.0) | 89 (59.3) | **0.039** |
| Previous mRS, n (%) |  |  | 0.084 |
| mRS 0 | 316 (80.2) | 107 (71.3) |  |
| mRS 1 | 45 (11.4) | 25 (16.7) |  |
| mRS 2 | 33 ( 8.4) | 18 (12.0) |  |
| Tobacco, n (%) |  |  | **0.025** |
| Non smoker | 262 (67.4) | 116 (78.9) |  |
| Active smoker | 72 (18.5) | 20 (13.6) |  |
| Past smoker | 55 (14.1) | 11 (7.5) |  |
| Alcohol, n (%) |  |  | 0.273 |
| Non Alcohol Use | 332 (86.5) | 133 (90.5) |  |
| Alcohol Use | 37 ( 9.6) | 12 (8.2) |  |
| Past Alcohol Use | 15 ( 3.9) | 2 (1.4) |  |
| Hypertension, n (%) | 256 (65.3) | 111 (74.0) | 0.067 |
| Dyslipidemia, n (%) | 235 (59.9) | 87 (58.4) | 0.816 |
| Diabetes, n (%) | 88 (22.3) | 49 (32.7) | **0.018** |
| Atrial Fibrillation, n (%) | 92 (26.6) | 41 (31.5) | 0.338 |
| Metallic Valve, n (%) | 15 ( 3.8) | 7 (4.7) | 0.830 |
| Previous Ischemic Stroke, n (%) | 55 (14.0) | 19 (12.7) | 0.784 |
| Previous Hematoma, n (%) | 5 ( 1.3) | 2 (1.3) | 1.000 |
| Previous Myocardial Infarction, n (%) | 42 (10.7) | 16 (10.7) | 1.000 |

**Table 2.** Baseline of significant clinical variables of ischemic stroke cohort (n=544). Results are expressed in mean (SD) or number of cases (%). P-value in bold are significant (p<0.05).

| n | Post-Stroke Infection in Ischemic Stroke Cohort |  |  |
| --- | --- | --- | --- |
|  | No<br>394 | Yes<br>150 | p |
| Admission Blood Glucose, mean (SD) | 124.57 (36.81) | 140.31 (53.99) | **<0.001** |
| Admission NIHSS, mean (SD) | 10.14 (7.70) | 17.21 (7.01) | **<0.001** |
| SU NIHSS, mean (SD) | 4.76 (5.70) | 12.39 (7.81) | **<0.001** |
| Infarct Volume (cc), mean (SD) | 21.13 (57.51) | 55.86 (78.23) | **<0.001** |
| Intra-arterial Occlusion, n (%) |  |  | **<0.001** |
| MCA M1 | 120 (36.6) | 61 (42.7) |  |
| MCA M2 | 66 (20.1) | 30 (21.0) |  |
| Carotid T | 39 (11.9) | 30 (21.0) |  |
| Basilar | 7 (2.1) | 3 (2.1) |  |
| Vertebral | 6 (1.8) | 0 (0.0) |  |
| PCA | 21 (6.4) | 3 (2.1) |  |
| ACA | 0 (0.0) | 3 (2.1) |  |
| Others/more | 69 (21.0) | 13 (9.1) |  |
| Thrombectomy, n (%) | 209 (55.0) | 125 (84.5) | **<0.001** |
| tPA, n (%) | 136 (35.9) | 57 (38.5) | 0.644 |
| Nasogastric Tube, n (%) | 8 (2.4) | 40 (27.2) | **<0.001** |
| Urinary Catheter, n (%) | 41 (12.2) | 66 (44.9) | **<0.001** |
| Intubation, n (%) | 3 (0.9) | 17 (11.6) | **<0.001** |
| 3 months Prognosis (mRS), n (%) |  |  | **<0.001** |
| Independent | 252 (72.4) | 50 (37.0) |  |
| Dependent | 76 (21.8) | 45 (33.3) |  |
| All-cause mortality | 20 (5.7) | 40 (29.6) |  |

### Blood samples processing and analysis by enzyme-linked immunosorbent assay (ELISA) and Multiplex

Platelet-poor plasma was obtained from blood samples. For neutrophil-specific NE and MPO a commercially available ELISA was used according to manufacturers’ instructions (Human PMN elastase ELISA kit, Abcam, Cambridge, MA, USA; Human MPO ELISA kit, R&D Systems, Minneapolis, MS, USA).

To study the systemic/acute immune response after stroke, we performed a commercial bead-based multiplex assay (LEGENDplex^TM^ Human Essential Immune Response Panel, BioLegend, San Diego, CA, USA) which allows us for simultaneous quantification of some key targets essential for immune response such as IL-4, IL-2, CXCL10, IL-1BETA, TNFALPHA, CCL2, IL-6, IL-10.

### Statistical analysis

Statistical analyses were performed using R software (v4.2.2, R Development Core Team). Continuous variables were assessed for normality with the Kolmogorov-Smirnov test and visual inspection (histograms and QQ-plots). Non-normal variables, including NE and Neutrophil-to-Lymphocyte ratio (NLR), were log-transformed to reduce skewness. Group differences were evaluated with independent t-tests for continuous variables and χ2-tests for categorical variables, with significance set at p<0.05. Missing data was treated as missing, potentially reducing the sample size in some models.

Multiple logistic regression analysis (MLRA) was performed using the stepwise Backward Wald method to identify independent predictors of post-stroke infection. Multicollinearity was assessed via the Variance Inflation Factor (VIF), confirming no significant collinearity (VIF <5 for all predictors). Receiver operating characteristic (ROC) curve analysis assessed model discrimination across adjustments.

Internal validation was conducted using propensity score matching (PSM) to minimize confounding (age, sex, CVRF), applying a 1:1 nearest neighbor method. All analyses were performed using R packages, including *stats, car, MatchIt, pROC, dplyr, tidyverse and ggplot2*.

## RESULTS

### 1- Characteristics and temporality of the different types of post-stroke infections

After carefully selecting patients based on inclusion and exclusion criteria, we included a total of 544 ischemic stroke patients in our study (Figure 1A). Of these, 150 (27.57%) suffered at least one infection during their hospitalization following the ischemic stroke (Table 1).

**Figure 1.**
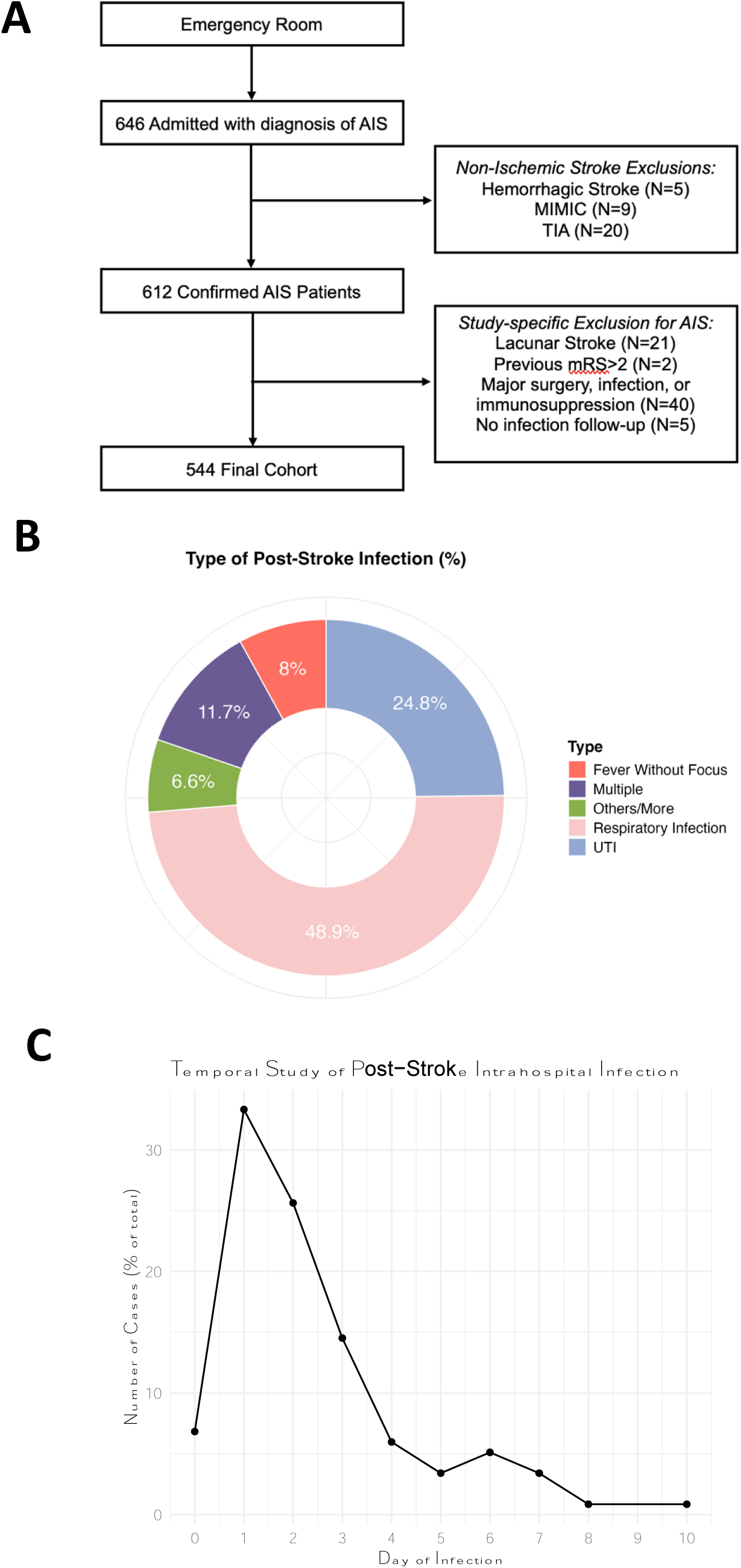
**A**. Flow diagram of patient’s recruitment. **B**. Frequency and type of infection in the cohort (UTI: urinary tract infection). **C**. Temporality of hospital admission of patients with ischemic stroke infection.

The most common infections were respiratory tract infections (48.9%), followed by urinary tract infections (24.8%) (Figure 1B). Analyzing the onset of the first infection symptoms (presence of fever, cough with imaging evidence of lung consolidation, dysuria, redness/pain at catheter site, etc), we found that 80% of cases occurred within the first three days after the stroke (Figure 1C).

Patients who developed infection display several significant baseline differences. As shown in Table 1, regarding demographic and the clinical characteristics, infected patients were significantly older, were more frequently women and diabetic. The rest of the clinical variables were included in Table 2. Regarding these variables, patients who developed infections had higher median NIHSS upon admission and at the stroke unit, this is 12-24 hours following admission. Median ASPECTS punctuation (more areas affected in MCA strokes) was lower in patients with infection. Infarct volume measured with admission MRI or control CT scan was bigger in patients with infection. Large vessel occlusion (MCA. M1, M2) was more frequent in infected patients. No differences were observed regarding the underlying etiology of stroke. Infected patients underwent mechanical thrombectomy more frequently, but rates of fibrinolysis with t-PA were similar in both groups. Regarding procedures associated with complications during patient hospitalization, a significant increase was observed in patients who subsequently developed infections. Specifically, there was a higher incidence of nasogastric tube placements, urinary catheterizations, and intubations. At 90 days, mRS and all-cause mortality were higher in patients who had suffered infection.

With respect to the laboratory variables, we found that there were notable differences between patients who would develop infections and those who would not (Table 3). Thus, total leukocytes were higher in the group of those who would become infected, both in admission and at the Stroke Unit, and we observe the same significant result in neutrophils, in admission and at Stroke Unit. Conversely, we see a reduction in lymphocytes in admission and at Stroke Unit. In parallel with the data observed for neutrophils, we saw an increase in NE and MPO levels in admission in those patients who would develop infections. Notably, since 80% of all infections occur within the first three days (Figure 1C), we focused our subsequent studies on this critical period.

**Table 3.** Clinical variables in the blood of stroke patients on admission and in the stroke unit at 12-24 hours. Results are expressed in mean (SD), p-value in bold are significant (p<0.05).

| n | Post-Stroke Infection in Ischemic Stroke Cohort |  |  |
| --- | --- | --- | --- |
|  | No<br>394 | Yes<br>150 | p |
| Admission Leukocytes, mean (SD) | 8.40 (2.69) | 9.39 (3.10) | **<0.001** |
| Admission Neutrophils, mean (SD) | 5.78 (2.54) | 6.94 (3.14) | **<0.001** |
| Admission Lymphocytes, mean (SD) | 1.82 (0.95) | 1.61 (0.89) | **0.021** |
| Admission Monocytes, mean (SD) | 0.67 (0.48) | 0.66 (0.27) | 0.804 |
| Admission Platelets, mean (SD) | 222.35 (69.81) | 226.13 (84.15) | 0.604 |
| Admission MPV, mean (SD) | 8.99 (1.08) | 9.23 (1.05) | 0.054 |
| Admission Neutrophil-Lymphocyte Ratio, mean (SD) | 4.28 (3.58) | 6.26 (5.94) | **<0.001** |
| SU Leukocytes, mean (SD) | 7.94 (2.52) | 9.46 (2.90) | **<0.001** |
| SU Neutrophils, mean (SD) | 5.65 (2.23) | 7.37 (2.74) | **<0.001** |
| SU Lymphocytes, mean (SD) | 1.54 (0.75) | 1.35 (0.97) | **0.022** |
| SU Monocytes, mean (SD) | 0.67 (0.60) | 0.80 (0.56) | **0.023** |
| SU Platelets, mean (SD) | 208.13 (69.13) | 201.86 (70.75) | 0.377 |
| Admission cell-free DNA, mean (SD) | 792.08 (134.26) | 788.15 (147.50) | 0.828 |
| Admission Neutrophil-Specific Elastase, mean (SD) | 43.90 (30.34) | 58.67 (46.12) | **<0.001** |
| SU Neutrophil-Specific Elastase, mean (SD) | 51.23 (75.92) | 59.56 (42.87) | 0.346 |
| Neutrophil-Specific Elastase Variation (%) (24h-admission), mean (SD) | 69.33 (395.62) | 39.20 (151.11) | 0.508 |
| Admission Myeloperoxidase, mean (SD) | 75.34 (78.99) | 96.58 (80.01) | **0.011** |
| Admission Blood Glucose, mean (SD) | 124.57 (36.81) | 140.31 (53.99) | **<0.001** |

### 2- Patients with a stroke-associated infection had higher pre-existing levels of neutrophil elastase

MLRA was performed for predictors of infection, based on a stepwise MLRA (Backward Wald, Figure 2A). Due to missing values, MLRA was performed on 474 of 544 patients. Following variables were included: NIHSS, age, sex, diabetes, NE (log), leukocyte, neutrophil counts, as well as the rest of the significant variables in the univariate analysis. Finally, in MLRA, admission NIHSS (OR: 1.12; 95% CI: 1.08-1.16), diabetes (OR: 2.05; 95% CI: 1.12-3.75) and NE (log) (OR: 2.15; 95% CI: 1.37-3.39) were independent predictors of infection (Adjustment B; Figure 2B) showing the strongest predictive performance with an AUC of 0.785 in ROC curve analysis. Comparatively, Adjustment A (NIHSS and diabetes) and Adjustment C (NIHSS, diabetes, and infarct volume) yielded AUCs of 0.756 and 0.779, respectively (Figure 2B). These findings suggest that NE values remain independently associated with infection risk, even after adjustment for diabetes and NIHSS scores, unlike infarct volume. The number of observations varied across models due to incomplete laboratory data for all the patients.

**Figure 2.**
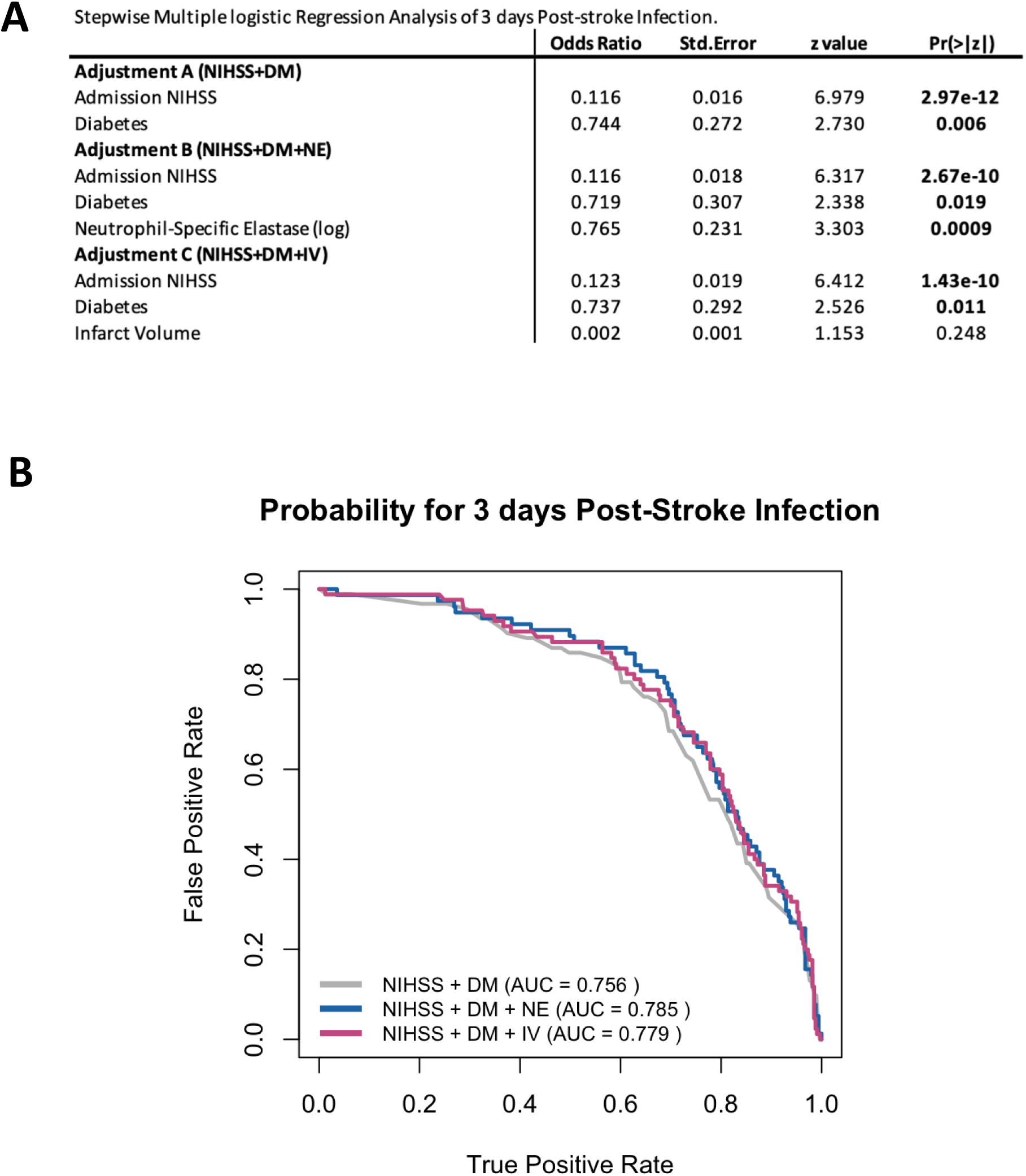
A. Stepwise multiple logistic regression analysis of three days post-stroke infections. Analysis was performed for predictors of post-stroke infection. **B**. Three adjustments and ROC curve analysis are shown.

### 3- Role of NE levels in predicting infections

To study the role of NE levels in predicting infection, a nested matched case-control analysis was performed to eliminate residual confounding (Figure 3A). Cases were defined as patients with post-stroke infections. Each case was assigned at least one matched control based on relevant clinical variables (age, sex, active cigarette smoking, alcohol use, NIHSS, diabetes, hypertension, dyslipidemia, auricular fibrillation, presence of a mechanical heart valve, previous ischemic stroke, or myocardial infarction, peripheral arteriopathy, chronic kidney disease and previous medication) by nearest neighbor matching method 1:1. This analysis yielded 102 cases and 106 matched controls. Matching was successful as cases and controls did not display significant changes in the selected variables. After successful matching, median levels of NE were still significantly higher in the case group than in the matched control group (Figure 3B).

**Figure 3.**
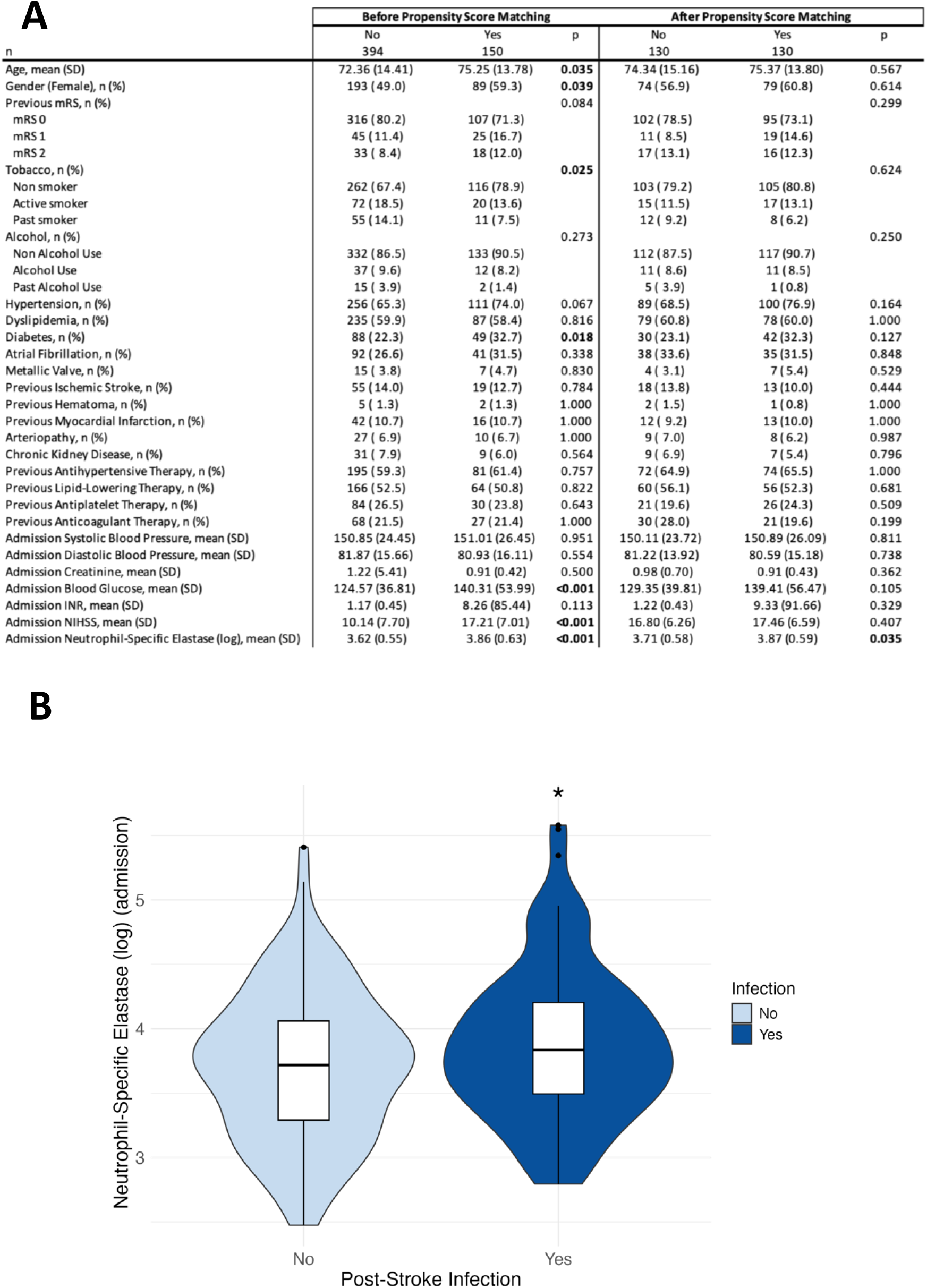
**A**. Nested matched case-control analysis. An internal validation was conducted using propensity score matching to minimize confounding, applying a 1:1 nearest neighbor method. **B**. After successful matching, median levels of NE (log) were analyzed by t-test. Significant p-value (p<0.05).

### 4- Patients with post-stroke infections had a pre-existing altered systemic immune profile

After verifying the importance of NE as a predictor of post-stroke infections, we wanted to study the immune response profile that patients would have at the time of admission and the next day in the stroke unit. The analysis of the immune profile was conducted by ensuring that patients included in the assay continued to meet the criteria for the nested matched analysis (Figure 3A). To this end, we analyzed the expression of different cytokines and chemokines in 74 patients (36 who did not develop infection and 38 who did) randomly selected from the patients included in the nested matched case-control analysis (Figure 4).

**Figure 4.**
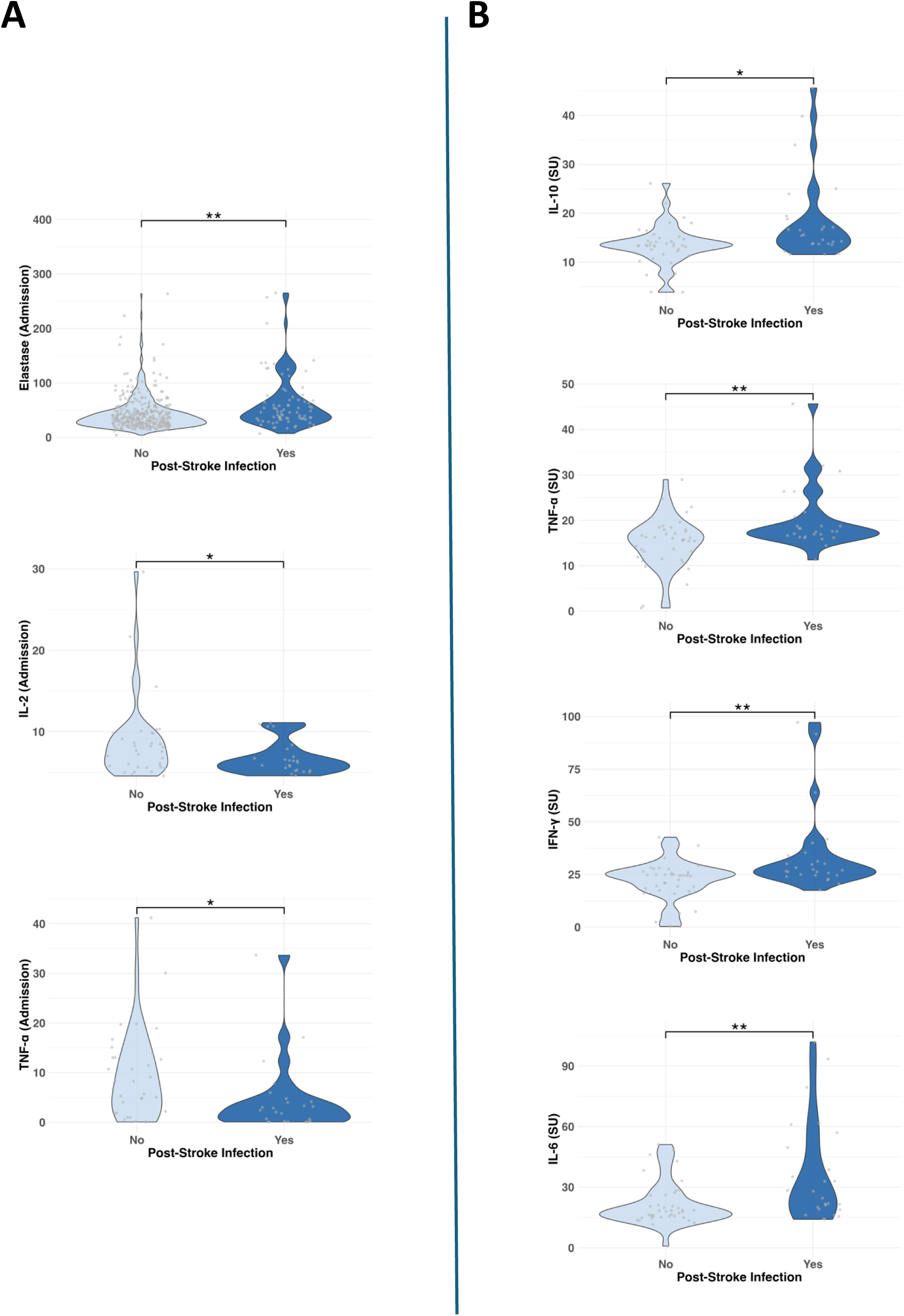
Immune profile in patients who develop early infection (day 1-3). Main differences in immune markers in the admission (**A**) and at 24h in the Stroke Unit (**B**).

The analysis shows that after admission, in addition to the increase in NE, patients who will become infected showed a reduction in the cytokines IL-2 and TNFα (Figure 4A). In contrast, at 24 hours, patients who will develop infections showed a marked pro- and anti-inflammatory profile, with a significant increase in the expression of the cytokines TNFα, IL-6, IL-10 and IFNγ (Figure 4B). Our results show that the immune profile for predicting the occurrence of infections is more accurate if they develop early (0 to 3 days after the ischemic stroke vs the whole period (data not shown).

To further focus the study, we analyzed whether any of the cytokines studied were related to a particular type of infection and found that both NE (admission and 24 hours) and IL-6 (24 hours) showed greater differences in respiratory infections (Figure 5).

**Figure 5.**
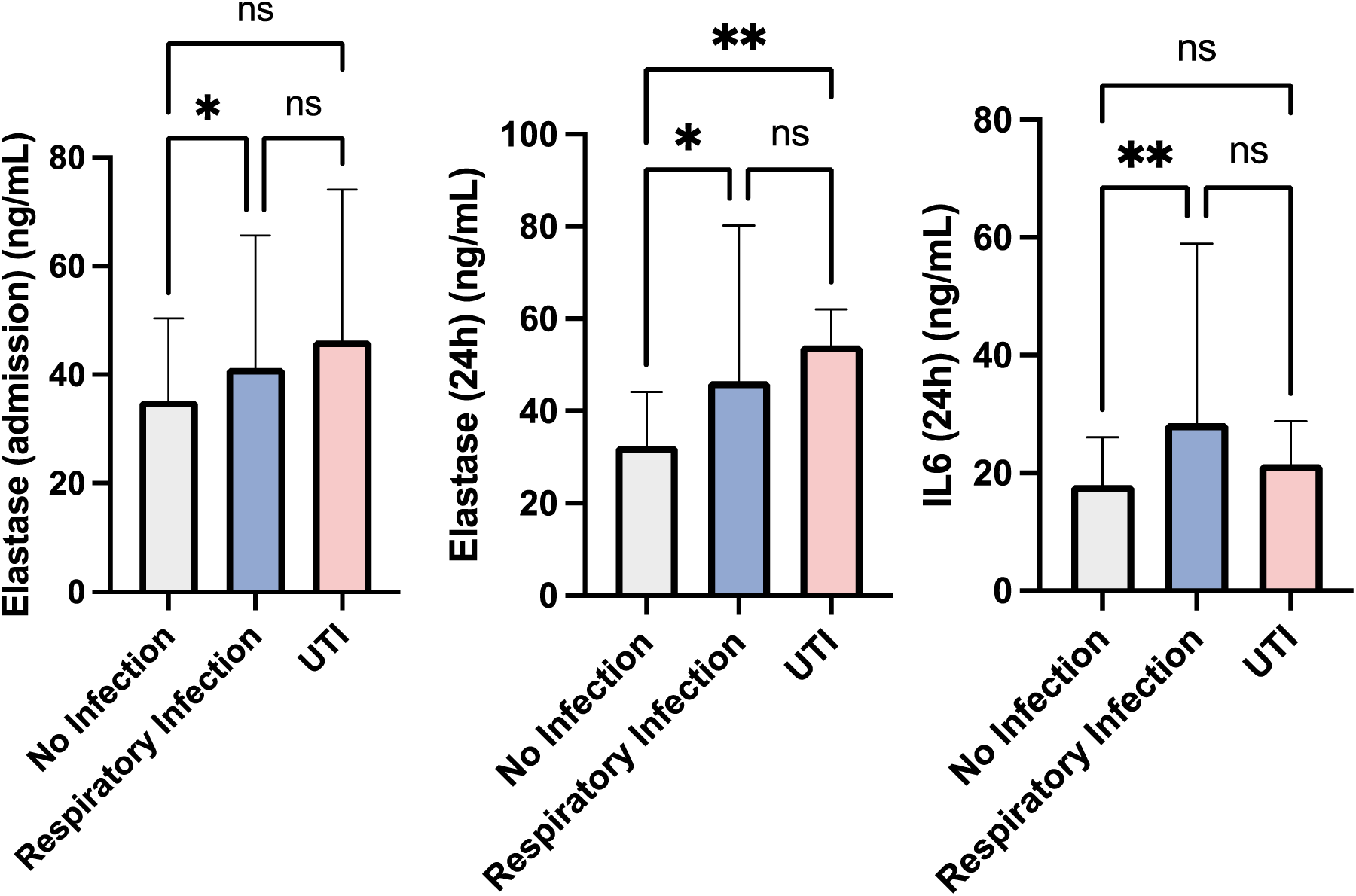
Differences in immune markers in the admission and at 24h in the Stroke Unit of the different types of infections (74 patients). \**P <* 0.05, \*\**P <* 0.01.

## DISCUSSION

Our study demonstrates for the first time that NE levels upon hospital admission are strongly associated with the development of post-stroke infections during hospitalization. Elevated NE levels were observed in these patients, raising the question of whether this increase reflects pre-existing chronic elevations or is a direct consequence of stroke-induced inflammation.

It is a well-established fact that previous risk factors increase the chances of suffering a stroke and developing more complications. Ischemic stroke predominantly affects individuals of advanced age, a group particularly susceptible to different inflammatory conditions due to diminished immune responses. This phenomenon, known as immunosenescence or inflamm-aging, involves chronic low-level inflammation resulting from an imbalance between pro-inflammatory and anti-inflammatory factors.^17^ On the other hand, it is well known that neutrophils in older adults have a weakened ability to eliminate bacteria, which further increases the risk of post-stroke infections.^18^ Our findings confirm that older individuals are more prone to infections. Thus, the decline in immune system efficiency and neutrophil function with aging may explain the link between age and disease severity.^19,20^

Diabetes and hyperglycemia are two of the most common risk factors linked to poor outcomes in ischemic stroke patients. Our results reveal that patients who developed infections were more likely to have diabetes and hyperglycemia. These factors can induce NETosis^20–22^ contributing to diabetes complications^23^ and exacerbating ischemic brain damage through hyperglycemia.^24^ Importantly, our matching analysis showed that NE levels remained higher in patients who developed infections, even when diabetes was not significantly different between non-infected and infected patients. Altogether, these results suggest that although diabetes has been reported as an important NETosis trigger, it was not the main cause of elevated NE levels in post-stroke infected patients.

We also observed that post-stroke infections are more common in women than in men. Key risk factors that differ by sex include diabetes and hypertension.^25^ Notably, diabetes is more strongly associated with the incidence of ischemic stroke in women than in men, with type 1 diabetes showing an even more pronounced difference.^26^ In addition to risk factors shared with men, such as hypertension and diabetes, women face unique risk factors, including early menopause and hormonal influences,^27^ which contribute to higher mortality rates and greater disability following a stroke.^28^

Despite the fundamental importance of NE in innate immunity, whose primary physiological function is to degrade phagocytosed materials such as elastin, collagen, and fibronectin,^29^ excessive activation of neutrophils under pathological conditions could lead to NE overactivation. This, in turn, may contribute to uncontrolled extracellular matrix degradation and tissue damage. Due to the large number of risk factors for stroke, it is difficult to determine the origin of increased NE levels. As previously discussed, certain risk factors like diabetes and age are known to exacerbate the mechanisms that can create a pro-inflammatory and pro-thrombotic environment. Over time, this imbalance may lead to persistent high NE levels,^30^ which contributes to further tissue damage and increases the risk of subsequent infections.^31^

NE has been linked to the pathogenesis of various diseases that affect the respiratory system, including pneumonia, acute lung injury, cystic fibrosis, and chronic obstructive pulmonary disease,^32^ in fact, NE’s role in pulmonary bacterial infections is well-documented, with studies proposing it as a biomarker.^33^ As our results show, the greatest increase in NE has been observed in respiratory infections which are more frequent and better predicted than other infections. The lungs are particularly susceptible to neutrophil-mediated damage due to their extensive capillary network, which facilitates significant neutrophil migration. Even in healthy individuals, neutrophil concentrations in the lungs are higher than in large blood vessels due to the low deformability of neutrophils that will keep them retained in the pulmonary microvasculature.^34^ This high density predisposes the lungs to tissue injury during excessive neutrophil activation.^35^

The level of NE expression is a critical determinant of immune response. Dysregulated NE activity not only damages tissues but also exacerbates inflammation by increasing reactive oxygen species and cytokine expression, inducing changes in neutrophil phenotypes, and inhibiting epithelial repair mechanisms.^32^ Our data show that there is an increase in cytokine release at 24 hours. In line with our results, in literature there are studies that highlight the importance of infections after stroke as independent predictors, such as the anti-inflammatory cytokines IL-10^2,36^ and IL-6.^37,38^ Furthermore, IL-6 has been shown to promote emergency granulopoiesis in response to systemic bacterial infections.^39^ This is one of the hypotheses that we are considering. During severe systemic infections, the high consumption of neutrophils to combat infection creates stress on the hematopoietic system which triggers a shift from steady state granulopoiesis to emergency granulopoiesis to meet the increased demand. Similarly, after the acute phase of a stroke, hyperactivation of the immune system may deplete mature neutrophils capable of degranulation and NETosis, potentially reducing the body’s ability to respond to infections, though this cannot be confirmed with our data. Another limitation of our study is the lack of comparative analyses between samples taken at admission and during peak infection-related fever. Such analyses could provide further insights into the role of coagulation and neutrophil dysfunction in the development of post-stroke infections. Future studies should aim to address this gap.

Our findings suggest that NE levels upon hospital admission may serve as a valuable biomarker for early identification of patients at high risk for respiratory infections following ischemic stroke. When combined with other risk factors, such as age and diabetes, these levels could improve predictive accuracy, enabling more targeted and effective treatment. Several clinical trials of anti-NE treatments for various respiratory syndromes (for review^40^) may also offer potential for treating conditions linked to excessive NE activity, such as post-stroke infections. Additionally, changes in key immune response molecules further support the identification of patients at higher risk for nosocomial infections, making prophylactic antibiotic therapy a consideration.

## CONTRIBUTIONS

Calleja, Martínez-Salio, Moro, Moraga and Lizasoain designed the research studies. Díaz-Benito, Alzamora, Ruiz, Ostos, Gutierrez-Sánchez, García-Culebras acquired the data. Díaz-Benito, Calleja, Alzamora, García-Culebras, Moro, Moraga and Lizasoain contributed to the analysis and interpretation of the results. Diaz-Benito, Calleja, Moro, Moraga and Lizasoain wrote the article, which all authors reviewed and approved.

## Data Availability

All data referred to in the manuscript are available upon request

## Non-standard Abbreviations and Acronyms

CVRF: cardiovascular risk factors
MCA: Middle cerebral artery
MLRA: Multiple logistic regression analysis
MPO: myeloperoxidase
mRS: modified Rankin Scale
NE: neutrophil elastase
NETs: Neutrophil extracellular traps
NIHSS: National Institutes of Health Stroke Scale

